# SARS-CoV-2 specific T cell responses are lower in children and increase with age and time after infection

**DOI:** 10.1101/2021.02.02.21250988

**Authors:** Carolyn A Cohen, Athena PY Li, Asmaa Hachim, David SC Hui, Mike YW Kwan, Owen TY Tsang, Susan S Chiu, Wai Hung Chan, Yat Sun Yau, Niloufar Kavian, Fionn NL Ma, Eric HY Lau, Samuel MS Cheng, Leo LM Poon, JS Malik Peiris, Sophie A Valkenburg

**Author notes:** Corresponding author: Dr Sophie A Valkenburg, HKU Pasteur Research Pole, School of Public Health, The University of Hong Kong.

## Abstract

SARS-CoV-2 infection of children leads to a mild illness and the immunological differences with adults remains unclear. We quantified the SARS-CoV-2 specific T cell responses in adults and children (<13 years of age) with RT-PCR confirmed asymptomatic and symptomatic infection for long-term memory, phenotype and polyfunctional cytokines. Acute and memory CD4^+^ T cell responses to structural SARS-CoV-2 proteins significantly increased with age, whilst CD8^+^ T cell responses increased with time post infection. Infected children had significantly lower CD4^+^ and CD8^+^ T cell responses to SARS-CoV-2 structural and ORF1ab proteins compared to infected adults. SARS-CoV-2-specific CD8^+^ T cell responses were comparable in magnitude to uninfected negative adult controls. In infected adults CD4^+^ T cell specificity was skewed towards structural peptides, whilst children had increased contribution of ORF1ab responses. This may reflect differing T cell compartmentalisation for antigen processing during antigen exposure or lower recruitment of memory populations. T cell polyfunctional cytokine production was comparable between children and adults, but children had a lower proportion of SARS-CoV-2 CD4^+^ T cell effector memory. Compared to adults, children had significantly lower levels of antibodies to β-coronaviruses, indicating differing baseline immunity. Total T follicular helper responses was increased in children during acute infection indicating rapid co-ordination of the T and B cell responses. However total monocyte responses were reduced in children which may be reflective of differing levels of inflammation between children and adults. Therefore, reduced prior β-coronavirus immunity and reduced activation and recruitment of *de novo* responses in children may drive milder COVID-19 pathogenesis.

## Introduction

A lack of pre-existing SARS-CoV-2-specifc protective antibodies has led to the rapid global spread of the novel coronavirus, however the large majority of infections are reportedly asymptomatic or mild (Ing et al., 2020). Previous COVID-19 infection may protect from reinfection (Addetia et al., 2020, Lumley et al., 2020) and neutralizing antibodies are likely to play an important protective role (Chandrashekar et al., 2020). However, the emergence of variant strains (e.g. 501Y.V2) suggests the possibility of escape from previous neutralizing antibody (Greaney et al., 2020, Cele et al., 2021). Antibody based treatment of established infection has had minimal beneficial effect on clinical outcome in COVID-19 patients (Weinreich et al., 2020) and may lead to emergence of escape mutant variants (Kemp et al., 2020). Dysregulated innate immune responses, such as auto-interferon antibodies or delayed responsiveness has been reported in some severe COVID-19 cases but cannot account for the majority of severe infections (Blanco-Melo et al., 2020, Chen et al., 2020, Wang et al., 2020). Importantly, a coordinated cellular immune response has been key to clinical resolution of SARS-CoV-2 infection (Rydyznski Moderbacher et al., 2020).

Pre-existing cross-reactive antibodies elicited by exposure to endemic human common cold coronaviruses such as the related β-coronavirus OC43 and HKU-1, do not prevent infection with SARS-CoV-2 (Anderson et al., 2020, Edridge et al., 2020). Furthermore, pre-existing cross-reactive T cell immunity generated by common cold coronaviruses has also been detected in the majority of people (Tan et al., 2021), with epitope conservation mostly reported in the ORF1ab non-structural proteins (Grifoni et al., 2020), but SARS-CoV-2 cross-reactive T cell responses have also been detected despite lower (<67%) epitope homology (Mateus et al., 2020). Upon infection, T cell responses shift towards Spike and Nucleocapsid structural proteins (Le Bert et al., 2020, Mateus et al., 2020). However, cross-reactive CD4^+^ T cell responses have been reported as similar (Mateus et al., 2020) or lower avidity and may be associated with worsening clinical outcomes (Bacher et al., 2020). In animal models of re-infection, Spike-specific CD8^+^ T cell responses can compensate for inadequate antibody responses and may provide an immune correlate of protection (McMahan et al., 2020). The magnitude of ORF1ab specific SARS-CoV-2 T cell responses during infection of adults does not differ with symptom severity but does associate with reduced duration of illness (Le Bert et al., 2020). Therefore, determining the balance and specificity of SARS-CoV-2-specific T cell responses for structural, accessory and non-structural proteins may inform the COVID-19 response and pathogenesis.

Following mild COVID-19 infection SARS-CoV-2 specific memory B cells are established for at least 6 months with long-term stability that may be recruited upon reinfection (Rodda et al., 2021). T cells following SARS-CoV infection in 2003 have reassuringly been detected 17 years after infection (Le Bert et al., 2020). Robust adaptive antibody and T cell responses have been reported in symptomatic and asymptomatic SARS-CoV-2 infected adults (Long et al., 2020, Sekine et al., 2020). Although serum antibody response to the common cold coronaviruses maybe long lasting, reinfection is common one or more years after infection (Edridge et al., 2020). The severity of COVID-19 may be reduced by rapid and early recruitment of established immune responses (Thevarajan et al., 2020, Chan et al., 2020, Tosif et al., 2020). The early and rapid recruitment of T follicular helper (Tfh) cells (Juno et al., 2020) drives early antibody development (Thevarajan et al., 2020) by germinal B cell responses leading to increasing neutralising antibody titers, however increased disease severity is associated with higher viral loads and antibody titers (Le Bert et al., 2020). The magnitude of the acute T cell responses in Middle Eastern Respiratory Syndrome (MERS), a related β-coronavirus, is negatively associated with the magnitude of the CD4^+^ T cell response and the duration of illness and thus antigen loads (Mok et al., 2020, Zhao et al., 2017).

In a small family case study, children (n=3) exposed to their SARS-CoV-2 infected parents displayed a coordinated recruitment of total T cells and specific antibodies however infection was not able to be virologically confirmed (Tosif et al., 2020). Asymptomatic infection may represent a large proportion of SARS-CoV-2 infections, particularly in children. The immunological differences of cellular recruitment for children and adults has not been sufficiently characterised to determine the immunological basis of differing diseases severity and outcomes of COVID-19.

In Hong Kong, effective public health measures of track, trace, quarantine of returned travellers and testing of quarantined close contacts has led to the identification of RT-PCR confirmed asymptomatic infections, even in young children. In this study, we assessed the balance of specificity, memory phenotype, cytokine quality and longitudinal stability of SARS-CoV-2 T cell responses in children (aged 2-13 years old) and adults with asymptomatic or symptomatic infection to address the role of T cells in the pathogenesis of milder disease in children.

## Materials and Methods

### Study population and clinical samples

Our study used samples from 24 children and 45 adults with RT-PCR confirmed SARS-CoV-2 infection in Hong Kong (Table 1). The days after onset of symptoms (for symptomatic infections) and days after first RT-PCR confirmation (for asymptomatic infections) was noted. All symptomatic or asymptomatic RT-PCR confirmed infections were hospitalized. Heparinised blood was collected at hospital admission (range: 1-14 days post symptom onset and/or RT-PCR confirmed infection), at discharge (range: 6-60 days) and at regular intervals after discharge for convalescent and long-term memory (range: 61-180 days) (Figure 1A). We used samples from a total of 45 adults (mean±stdev: 43.1±13.7, range: 20-65 years) and 24 children (8.1±3.9, 1.92 (23 months)-13 years). We had 95 longitudinal samples from 46 subjects with 2 to 3 sampling time-points and 55 early acute time-points samples (< day 14) (Figure 1A). Samples of comparable time-points were used from children (32.5±40.4, 2-138 days) and adults (28.9±39.6, 1-180 days) (Table 1).

**Table 1 –.**
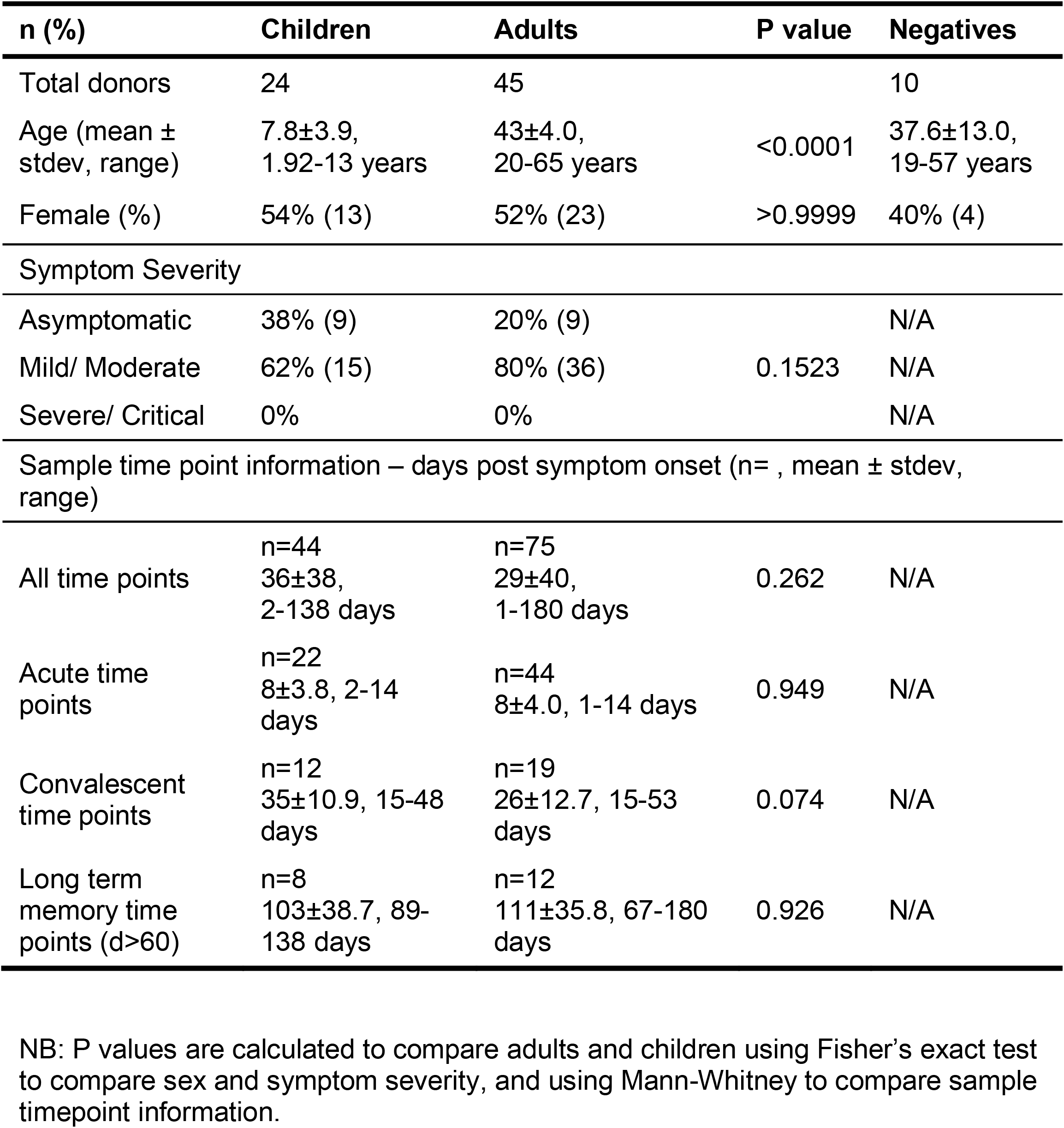
Summary of cohort information. Samples from SARS-CoV-2 infected children and adults, and negative controls forming a cohort where samples were used in multiple cellular and ELISA assays.

| n (%) | Children | Adults | P value | Negatives |
| --- | --- | --- | --- | --- |
| Total donors | 24 | 45 |  | 10 |
| Age (mean ± stdev, range) | 7.8±3.9, 1.92-13 years | 43±4.0, 20-65 years | <0.0001 | 37.6±13.0, 19-57 years |
| Female (%) | 54% (13) | 52% (23) | >0.9999 | 40% (4) |
| Symptom Severity |  |  |  |  |
| Asymptomatic | 38% (9) | 20% (9) |  | N/A |
| Mild/ Moderate | 62% (15) | 80% (36) | 0.1523 | N/A |
| Severe/ Critical | 0% | 0% |  | N/A |
| Sample time point information – days post symptom onset (n= , mean ± stdev, range) | | | | |
| All time points | n=44<br>36±38, 2-138 days | n=75<br>29±40, 1-180 days | 0.262 | N/A |
| Acute time points | n=22<br>8±3.8, 2-14 days | n=44<br>8±4.0, 1-14 days | 0.949 | N/A |
| Convalescent time points | n=12<br>35±10.9, 15-48 days | n=19<br>26±12.7, 15-53 days | 0.074 | N/A |
| Long term memory time points (d>60) | n=8<br>103±38.7, 89-138 days | n=12<br>111±35.8, 67-180 days | 0.926 | N/A |

NB: P values are calculated to compare adults and children using Fisher's exact test to compare sex and symptom severity, and using Mann-Whitney to compare sample timepoint information.

**Figure 1;.**
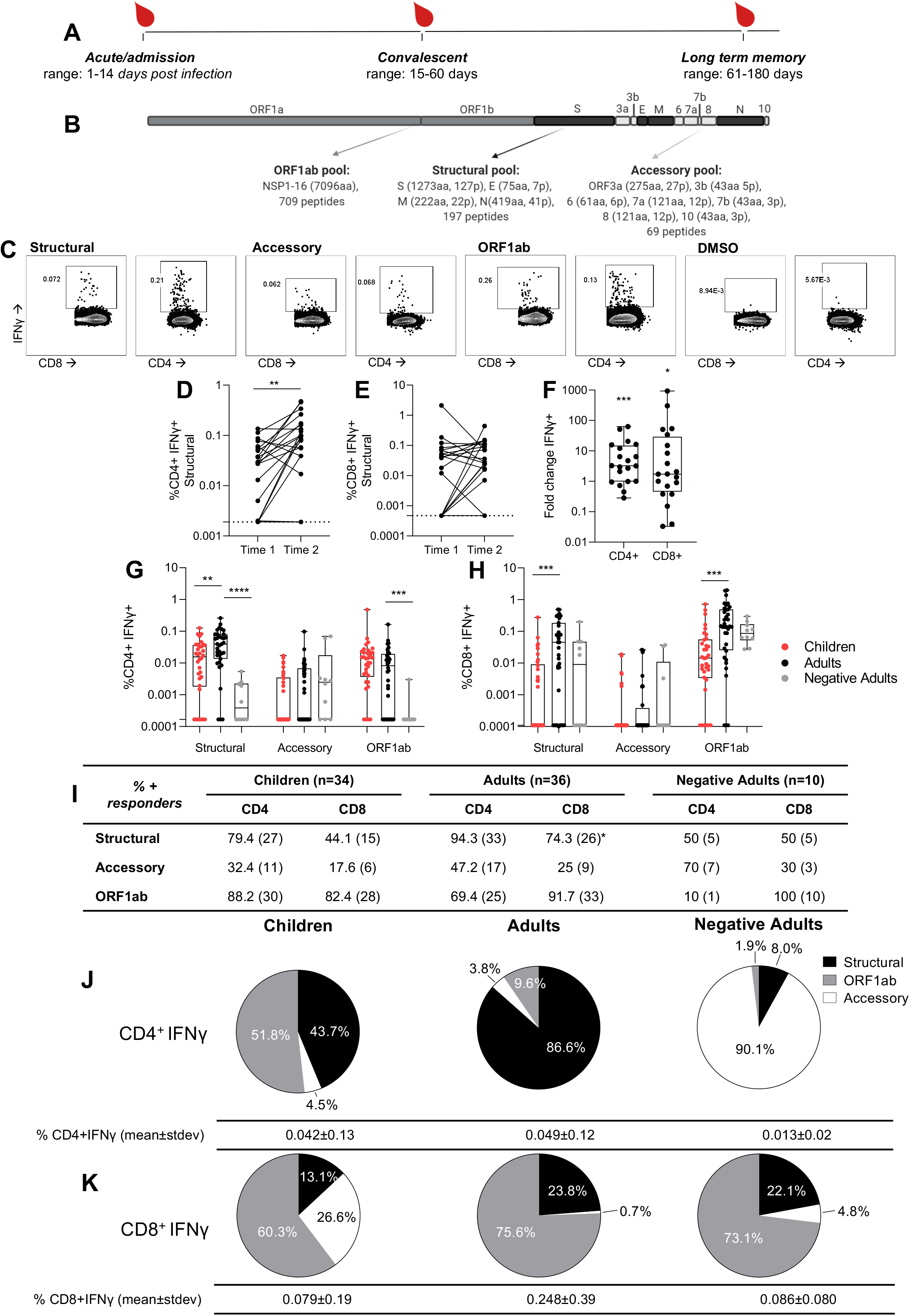
Children have lower CD4^+^ and CD8 ^+^ T cell responses than adults with COVID-19. CD8^+^ T cell responses are predominantly ORF1ab specific, while children and adults have different CD4^+^ T cells targets. (A) Heparinised blood samples for PBMCs were collected from COVID-19 patients in Hong Kong during the course of infection and recovery. (B) Overlapping peptide pools of the whole SARS-CoV-2 proteome were generated to represent ORF1ab, Structural, and Accessory proteins with amino acids (aa) and peptides (p) per protein shown. (C) PBMCs were stimulated with peptide pools or a DMSO control and IFNγ production of CD4^**+**^ and CD8^**+**^ T cells measured by flow cytometry. Paired time points at hospital admission and discharge (time 1: mean 7.25 +/- stdev 4.6 days post infection, range 3 to 18; time 2: mean 13.4 +/-stdev 4.4, range 6 to 21) for paired background (DMSO) subtracted structural specific IFNγ response of CD4^**+**^ (D) and CD8^**+**^ (E) T cells (n=20 adults). Wilcoxon test was used to determine differences **p<0.01. Dotted lines represent limit of detection following background subtraction (CD4=0.0019, CD8=0.00047). (F) The fold change of paired structural specific CD4^**+**^ and CD8^**+**^ T cells responses from (D, E), significance calculated using One sample Wilcoxon test against a theoretical median of 1. Dotted line at 1 indicates no fold change. The SARS- CoV-2 CD4^**+**^ (G) or CD8^**+**^ (H) T cell responses of COVID-19 children (n= 34), adults (n=36) (mean±stdev: 42±44, range 1-180 days) and negatives (n= 10). Data are displayed as individual responses to each peptide pool with IFNγ production to paired DMSO subtracted. The dotted line represents the lower limit of detection, determined as the smallest calculated value above the DMSO background response (IFNγ of CD4^+^=0.00017%, IFNγ of CD8^+^=0.00011%). Comparisons between groups were performed using Mann-Whitney test with the effect of the sampling time accounted for, statistical differences are indicated by *p<0.01, **p<0.001, ***p=0.0001. Values above the limit are used to classify participants as responders and presented as a percentage with the numbers of responders in brackets (I). Differences between children (n=34) and adults (n=36) from all time points (1 to 180 days post symptom onset) were determined by Fisher’s exact test and displayed in the adults column where *p<0.05. Pie charts show the proportion of total IFNγ^+^ CD4^**+**^ (J) and CD8^**+**^ (K) SARS-CoV-2 responses with DMSO subtracted in children (n=34), adults (n=36) and negatives (n=10) (from G, H). Values below the limit of detection assigned the value of 0.

The study was approved by the institutional review board of the respective hospitals, viz. Kowloon West Cluster (KW/EX-20-039 (144-27)), Kowloon Central / Kowloon East cluster (KC/KE-20-0154/ER2) and HKU/HA Hong Kong West Cluster (UW 20-273, UW20- 169), Joint Chinese University of Hong Kong-New Territories East Cluster Clinical Research Ethics Committee (CREC 2020.229). All of patients provided informed consent. The collection of SARS-CoV-2 seronegative adult negative control blood donors (37.6±13.0, 19-57 years) was approved by the Institutional Review Board of The Hong Kong University and the Hong Kong Island West Cluster of Hospitals (UW16-254).

Plasma was isolated, stored at -80°C and heat inactivated (HI) at 56°C for 30 minutes upon testing. Peripheral blood mononuclear cells (PBMC) were isolated by Ficoll-Paque (GE Healthcare) separation using Leucosep tubes (Greiner Bio-one) and cryopreserved in liquid nitrogen for batched analysis.

### SARS-CoV-2 overlapping peptide pools for T cell stimulation

An overlapping peptide library was made covering the whole SARS-CoV-2 proteome with 20 amino acid (aa) length and 10 aa overlap (Genscript). The amino acid sequence of the peptide pools was based on βCoV/Hong Kong/VM20001061/2020 strain (GISAID ID: EPI_ISL_412028). Peptides were dissolved in deionised water, 10% acetic acid, or DMSO according to their biochemical properties and charge. A pool of 197 peptides representing Structural proteins: from S (1273aa, 127 peptides), N (419aa, 41 peptides), E (75aa, 7 peptides), M (222aa, 22 peptides), with a DMSO concentration of 0.6%. The ORF1ab peptide pool consisted of 709 peptides for the NSP1-16 proteins (7096aa), with a DMSO concentration of 2.1%. An accessory peptide pool of 69 peptides for the ORF3a (275aa, 27 peptides), ORF3b (43aa, 5 peptides), ORF6 (61aa, 6 peptides), ORF7a (121aa, 12 peptides), ORF7b (43aa, 3 peptides), ORF8 (121aa, 12 peptides), ORF10 (43aa, 3 peptides) proteins with a DMSO concentration of 0.2% (Figure 1B). Experimental controls included: cytomegalovirus (CMV) peptide pool (Grifoni et al., 2020) and PMA/ionomycin as positive controls, and for negative controls media alone and average DMSO control (1.0% concentration) for background cytokine production (Supplementary Figure 1B). SARS-CoV-2 peptide Megapools (Spike plus all pool, 467 peptides) for predicted MHC restricted peptides covering all proteins of the genome for CD4^+^ T cells and CMV from Grifoni *et al*. were used as initial positive controls (Grifoni et al., 2020).

### SARS-CoV-2-specific T cells by Intracellular Cytokine Staining (ICS)

Cryopreserved PBMCs were thawed and re-stimulated with overlapping peptide pools representing the SARS-CoV-2 structural proteins, accessory proteins, or ORF1ab (300 nM), DMSO (1% in RPMI), CMV peptide pool, PMA/ionomycin (1% in PBS) or RPMI alone for 6 hours at 37°C. Golgi Plug (BD) containing Brefeldin A (1% in PBS), and Golgi Stop (BD) containing Monensin (0.67% in PBS) was added at 2 hours during stimulation. Cells were stained with Zombie-NIR (all antibodies from Biolegend and clone used) followed by anti-human CD3-PE/Dazzle 594 (UCHT1), CD4-BV605 (OKT4), CD8-AlexaFluor700 (SK1), CD107a-PacificBlue (H4A3), CCR5-PE (J418F1), CCR7-PerCP/Cy5.5 (G043H7) and CD45RA-APC (HI100) and a dump channel containing CD19-BV510 (HIB19), CD56- BV510 (HCD56) and CD14-BV510 (M5E2). Cells were then permeabilised and fixed (BD Cytofix/cytoperm) and further stained for anti-human IFNγ-FITC (4S.B3), IL-2-PECy7 (MQ1-17H12), TNF-α-BV711 (MAb11). Stained cells were acquired via flow cytometry (AttuneNxT) and analysed by FlowJo v10 (Supplementary figure 1). Experiments were repeated twice on independent samples.

### Immunostaining of Monocytes, T Follicular Helper (Tfh) cells and Plasmablasts

Whole blood samples were stained with an antibody panel (all Biolegend and clone used), and live/dead Zombie-NIR to identify monocytes, Tfh and plasmablast responses (Supplementary Figure 2). The combined monocytes/plasmablast panel contained: anti-human CD16-PE (3G8), CD14-PerCPCy5.5 (M5E2), HLA-DR-BV605 (L243), CCR2-APC (K035C2), CD19-BV510 (HIB19), CD27-FITC (M-T271) and CD38-BV421 (HIT2). The Tfh panel contained: anti-human CD4-AlexaFluor700 (SK3), CXCR5-PerCPCy5.5 (J252D4), CD45RA-FITC (HI100), CCR6-BV605 (G034E3), CXCR3-APC (G025H7), PD-1-BV711 (EH12.2H7) and ICOS-PE (C398.4A). Cells were acquired by flow cytometry (AttuneNxT).

**Figure 2:**
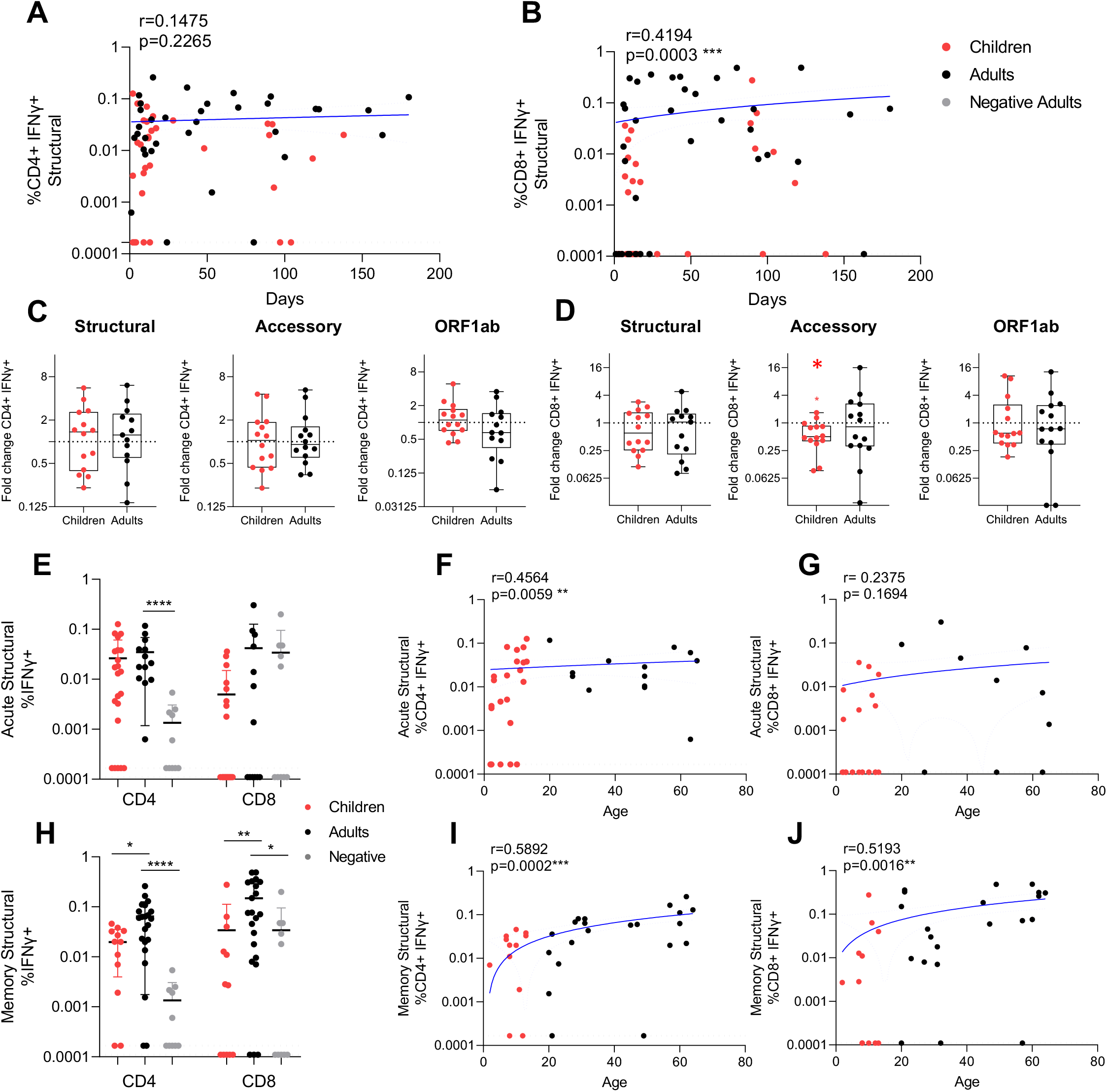
SARS-CoV-2 specific CD4+ and CD8+ T cell responses increase over time and age. Correlation of IFNγ responses for CD4^+^ (A) and CD8^+^ (B) T cells against the structural peptide pool with children (red) (n=34) and adults (black) (n=36) (with background IFNγ production to DMSO subtracted), against days post symptom onset. Black dotted lines represent the limit of detection (IFNγ of CD4^+^=0.000167 (A), IFNγ of CD8^+^=0.00011(B)). Fold change of IFNγ CD4^+^ (C) and CD8^+^ (D) T cell responses were calculated as the later time point (mean±stdev: 32.8±35.7 days, range: 9-138) over admission time point responses (mean±stdev: 7.6±4.2, range: 2-15)) in response to the structural, accessory and ORF1ab peptide pools in children and adults from two independent experiments (children n=14, adults n=14). One sample Wilcoxon tests were used for determining significance of fold changes, were *p<0.05. Acute (samples <14 days post symptom onset, mean±stdev: 8.0±3.8, range: 1-14, n=22 children, n=14 adults) (E-G), and convalescent/memory (H-J) (mean±stdev: 70.5±41.9, range: 15-180 days post symptom onset, n=12 children, n=22 adults) IFNγ structural specific (F, I) CD4^+^ and (G, J) CD8^+^ T cell responses and negative controls (n=10). For statistical comparisons between children and adults, or adults and negatives, Mann-Whitney tests were performed, *p<0.05, **p<0.01, ***p<0.001, ****p<0.0001. The magnitude of the acute (from E) and memory (from H) structural IFNγ CD4^+^ (F, I) and CD8^+^ (G, J) T cell response with age. R values are calculated using Spearman’s correlation and *p<0.05, **p<0.01, ***p<0.001, ****p<0.0001. Blue lines of linear regression represent the overall trend, and blue dotted lines show the upper and lower 95% confidence intervals. All data points are individual responses minus paired background IFNγ response to a DMSO control.

### Spikes-specific IgG quantification by Enzyme-linked immunosorbent assay (ELISA)

Plates (Nunc MaxiSorp, Thermofisher Scientific) were coated with one representative coronavirus Spike protein at a time. Plates were coated with either 80 ng/ml of purified baculovirus-expressed Spike protein from 229E, NL63, HKU-1 and OC43 (SinoBiological). Plates were rinsed, blocked with 1% FBS in PBS, incubated with 1:100 HI plasma diluted in 0.05% Tween-20/0.1% FBS in PBS for 2 hours then rinsed again, and incubated for 2 hours with IgG-HRP (1:5000, G8-185; BD). HRP was revealed by stabilized hydrogen peroxide and tetramethylbenzidine (R&D systems) for 20 minutes, stopped with 2N sulphuric acid and absorbance values were recorded at 450nm on a spectrophotometer (Tecan Life Sciences).

### Statistical Analysis

Statistical analysis was performed on Prism 9 (Graphpad). For two-way comparison, the Wilcoxon signed-rank test (paired) or Mann-Whitney t-test (unpaired) was used. For multiple-group comparisons, a Friedman (paired) or Kruskal-Wallis (unpaired) test, followed by the Dunn-Bonferroni post-hoc test was used. The One sample Wilcoxon signed-rank test was used for comparisons against a hypothetical value for fold changes. Correlations were performed using the Spearman’s test. To account for correlation due to multiple measurements from the same patients, a linear random effects model was fitted (Supplementary table 1). The model also tested the linear time trend by days after illness onset, and potential differences by age, sex and symptomatic patients. Differences were tested using Mann-Whitney test. Differences in baseline characteristics was detected with the chi square test. Adjusted *p* values < 0.05 were considered statistically significant.

## Data availability statement

The protein, peptide sequences and data that support the findings of this study are available from the corresponding author upon request. The amino acid sequence of the peptide pools was based on βCoV/Hong Kong/VM20001061/2020 strain (GISAID ID: EPI_ISL_412028). Data from flow cytometry and ELISA IgG responses with background subtracted are indicated, and representative flow cytometry plots are shown.

## Results

### Recent infection is associated with an increase of CD4^+^ T cell responses to structural proteins

SARS-CoV-2 specific T cell responses were assessed from COVID-19 cases in children and adults, and in adult negative controls. SARS-CoV-2 consists of 4 structural proteins, an extensive ORF1ab which encodes 16 non-structural proteins, and 7 accessory proteins. The relative expression of the structural proteins versus accessory and non-structural proteins during SARS-CoV-2 virus replication may impact their immunogenicity. Cross-reactivity with common cold viruses (Edridge et al., 2020) may also affect the magnitude of T cell responses elicited. Due to limited cell numbers of our samples, peptide or protein specific mapping was not possible. Therefore direct *ex vivo* CD4^+^ and CD8^+^ T cell responses were assessed for overlapping peptide pools of structural, accessory and ORF1ab proteins respectively, (Figure 1B) using IFNγ production, a key anti-viral cytokine as a read-out for specificity (Figure 1C). Paired samples from SARS-CoV-2 infected adults at hospital admission (time 1) and discharge (time 2) showed an increase in structural specific IFNγ^+^ CD4^+^ T cells (Figure 1DF, fold change p=0.0005) and to a lesser extent CD8^+^ T cells (Figure 1EF, fold change p=0.0230).

The magnitude of SARS-CoV-2 specific CD4^+^ (Figure 1E) and CD8^+^ (Figure 1F) T cells for structural, accessory and ORF1ab proteins was compared between adult patients versus adult negative controls to establish assay specificity and cross-reactivity. We then compared the T cell responses of the adult infections versus paediatric infections to define differences with age. The IFNγ^+^ CD4^+^ T cell responses towards structural proteins of SARS-CoV-2 were significantly increased in adults (mean±stdev: 0.0533±0.0549%), compared to both children (0.0240±0.0292%, p=0.0031) and adult negative controls (0.0013±0.0005%, p<0.01) (Figure 1G). The majority of infected adults (94.3%) mounted a structural-specific CD4^+^ T cell responses (above DMSO background) (Figure 1I), whilst only 79.4% of children and 50% of adult negative controls had such responses (Figure 1I). Despite the higher magnitude of responses to structural proteins in infected adults than children, the proportion of responders against each peptide pool was not significantly different between adults and children, except for structural CD8^+^ responses (Figure 1I). Therefore, the majority of our later analyses focusses on structural specific T cell responses.

The accessory-specific CD4^+^ T cell response was comparable in infected children, infected adults and adult negative controls (Figure 1G). In infected adults, the structural-protein-specific CD4^+^ T cell responses (86.6%) contributed most to the SARS-CoV-2 specific response (Figure 1J), than ORF1ab (9.6%) and accessory (3.8%) responses. By contrast, the SARS-CoV-2 specific response in infected children’s CD4^+^ T cell responses were dominated more by ORF1ab (51.8%) than structural specific responses (43.7%). Responses from adult negative controls that recognised SARS-CoV-2 peptides were predominately specific for accessory peptides (90.1%), however the total response was very low in magnitude, at only 0.013±0.02% of CD4^+^ T cells (Figure 1J).

Infected adults did not have significantly higher CD8^+^ T cell responses compared to adult negative controls (Figure 1H) indicating cross-reactivity and little amplification of SARS-CoV-2 CD8^+^ T cell responses by infection (Figure 1EF). But infected children had significantly reduced CD8^+^ T cell responses compared to infected adults for structural and ORF1ab responses (Figure 1H).

However baseline differences exist between adults and children for non-specific T cell activation (Lewis et al., 1991, Rudolph et al., 2018, Booth et al., 2014). The baseline activation (by DMSO) and overall maximum activation (by PMA/ionomycin) is lower in children, and responsiveness significantly increased with age (Supplementary Figure 2 A-D). Adult negative controls had comparable background IFNγ induction compared to infected adults (Supplementary Figure 2A), However, the maximum responses by PMA/ionomycin was lower in CD4^+^ T cells and higher in CD8^+^ T cells in negative control adults compared to infected adults (Supplementary Figure 2B), which may indicate T cell activation is refractory based on recent infection (Crawford et al., 2013). Normalisation of structural specific T cells by % of maximum PMA/ionomycin responses shows comparable responses across all groups (Supplementary Figure 2F). However the fine specificity of identifying low frequency antigen specific T cells is obscured, therefore T cell responses should be considered directly *ex vivo* with background subtracted not normalised for maximum activation. There was a significant correlation between PMA/ionomycin and structural specific CD4^+^ T cell IFNγ production (r=0.6384, p=0.0001, Supplementary Figure 2G), but not CD8^+^ T cell responses (r=0.2568, p=0.1707, Supplementary Figure 2H).

Furthermore, stratification of subjects for asymptomatic and symptomatic infection did not reveal any further significant differences for T cell response magnitude (Supplementary Figure 3AB) or contribution of peptide specificities (Supplementary Figure 3CD) between controls and COVID-19 adults and children.

**Figure 3.**
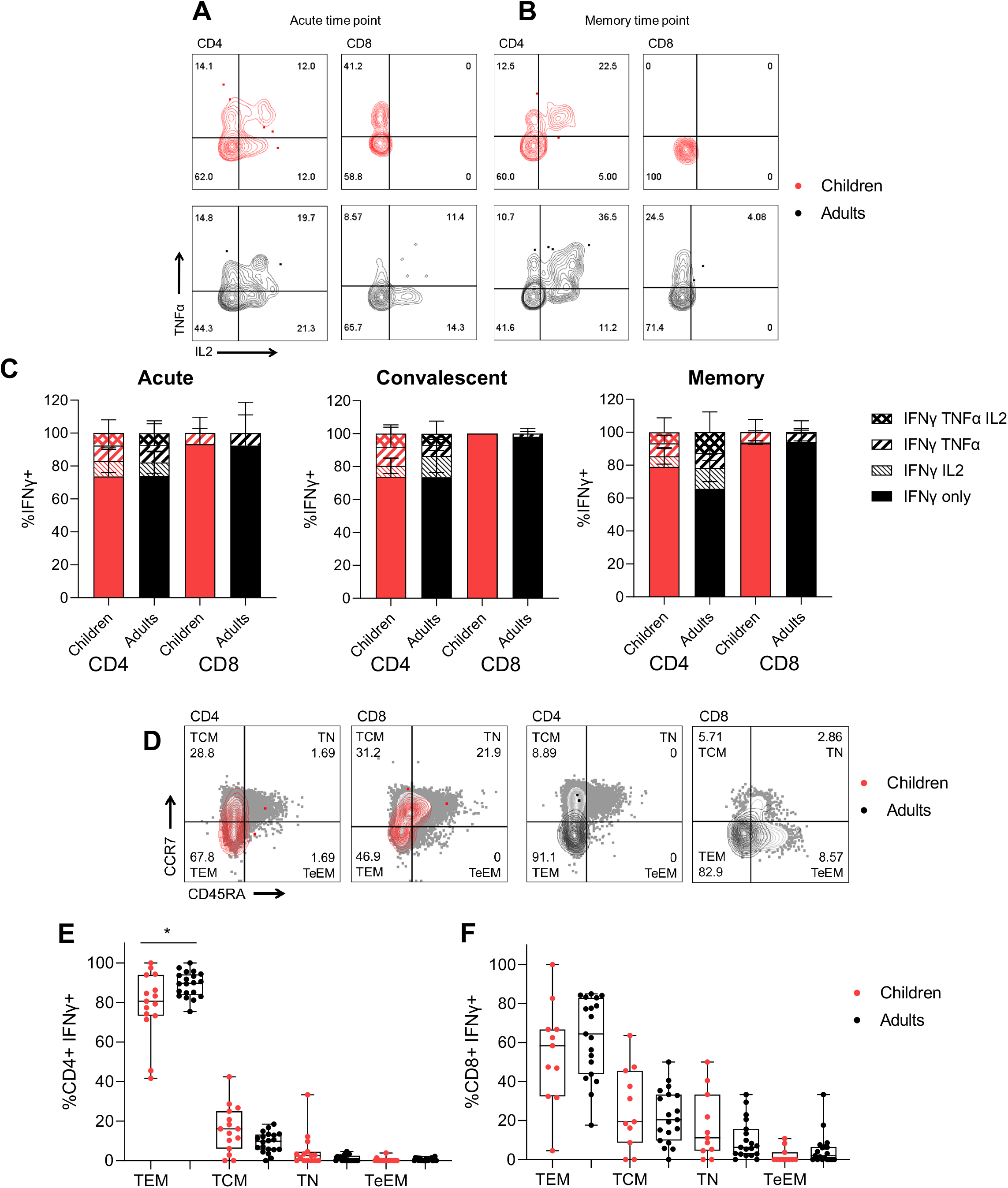
Cytokine polyfunctional quality is comparable in COVID-19 children and adults, whilst T effector memory phenotype is increased in adults. Representative FACS plots of TNFα and IL2 producing IFNγ^+^ CD4^+^ and CD8^+^ T cells of children (red) and adults (black) at acute (d<14) (A) and memory (child: 118 days, adult: 94 days) (B) time points. (C) The proportion of IFNγ producing CD4^+^ and CD8^+^ T cells which are single, double or triple cytokine producers at acute (<14 days), convalescent (15-60 days) or memory (61-180 days) time points post symptom onset. Kruskal Wallis test for multiple comparisons was carried out to compare each group between children and adults. (D) Representative FACS plots showing memory phenotypes of IFNγ^+^ CD4^+^ and CD8^+^ T cells based on expression of CCR7 and CD45RA. Sections are T effector memory (TEM), central memory (TCM), terminal effector memory (TeEM) or naïve (TN). Memory phenotype responses in IFNγ^+^ CD4^+^ (E) and CD8^+^ (F) T cells of responders at later time points (15-180 days post symptom onset). Comparisons between children (n=15) and adults (n=20) in each group was performed using Mann-Whitney test, *p<0.05.

### Recruitment of early cellular responses

Coordination of the early acute response to SARS-CoV-2 infection is important to drive innate responses (Zhang et al., 2021), and early antibody production (Thevarajan et al., 2020) for improved patient outcomes. Therefore, we assessed the recruitment and activation of monocytes, total Tfh cells and plasmablasts (also known as antibody producing cells) during acute (<14 days post infection) SARS-CoV-2 infection (Supplementary Figure 4). The total monocytes showed reduced responses in children compared to infected adults (Supplementary Figure 4B), furthermore children had reduced inflammatory type monocytes (Supplementary Figure 4C), where these have been found to also be elevated in COVID-19 patients, but lower in severe outcomes in adults (Zhang et al., 2021, Mann et al., 2020). Meanwhile, infected children and adults showed comparable levels of monocyte recruitment from bone marrow (CCR2) compared to infected adults (Supplementary Figure 4D).

**Figure 4.**
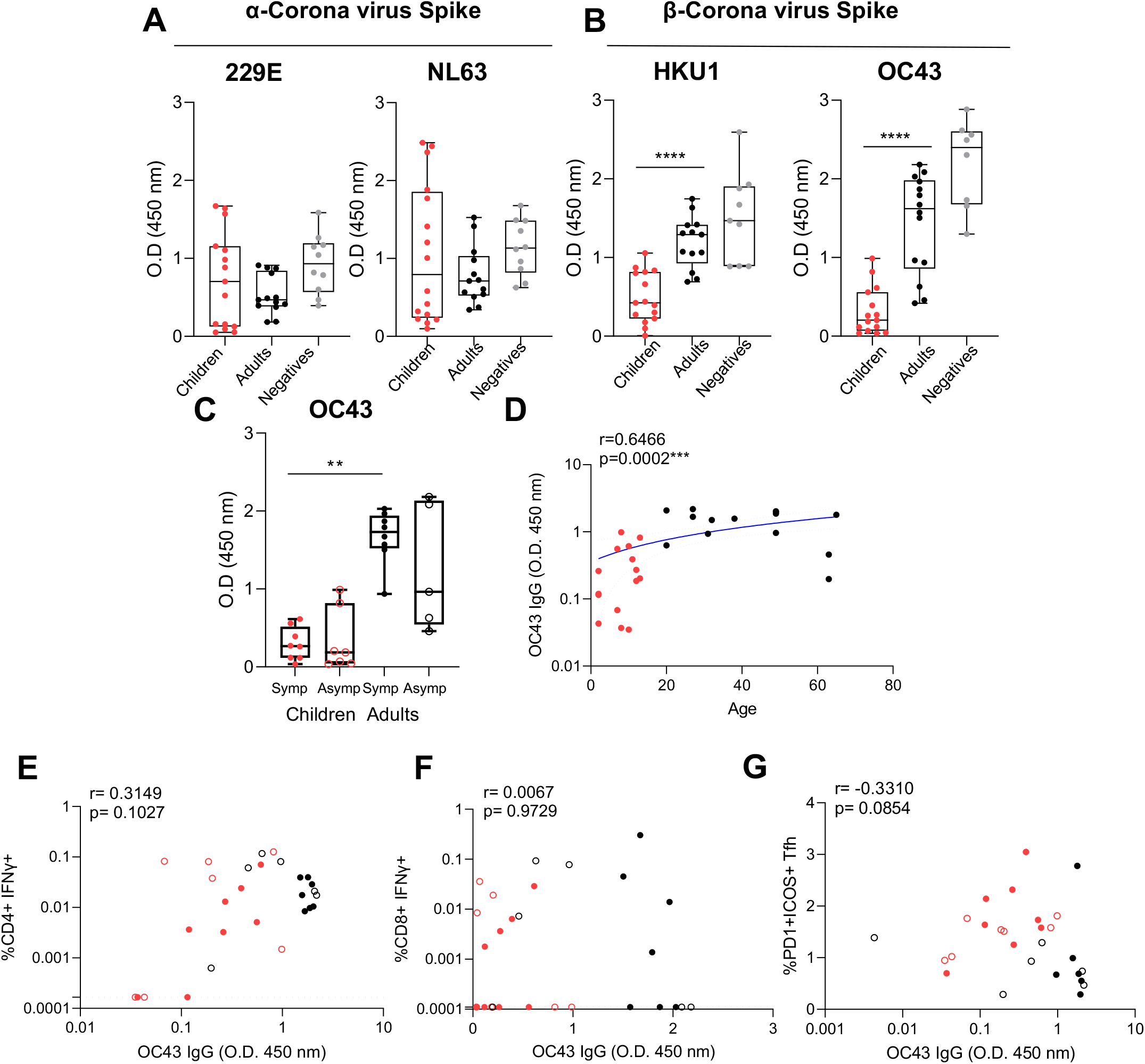
Previous exposure to common cold beta coronaviruses and T cell responses. Total IgG responses to the Spike protein (S1+S2) of common cold α (229E, NL63) (A) and β (HKU1, OC43) (B) coronaviruses measured by ELISA from acute time points (mean±stdev: 8±3.8, range: 2-14 days post infection). (C) Stratification of OC43 IgG response by symptomatic (closed circles, n=8 children, n=8 adults) and asymptomatic (open circles, n=8 children, n=5 adults). (A-C) Data is representative of individual donor responses with background subtracted (non-specific protein block), and displayed with the median, upper and lower quartiles and minimum and maximum. Comparison between children (n=15) and adults (n=14) or adults negative controls (n=10) was performed using Mann-Whitney test where **p<0.01, ***p<0.001, ****p<0.0001. (C) Multiple comparisons between symptomatic and asymptomatic adults and children were carried out using Kruskal Walis tests, where **p<0.01. (D) Correlation of OC43 IgG and age. A blue line of linear regression represents the overall trend, and blue dotted lines show the upper and lower 95% confidence intervals. Correlation of structural SARS-CoV-2 specific IFNγ^+^ CD4^+^ (E) or CD8^+^ (F) T cell responses and OC43 Spike IgG. (G) Correlation of activated Tfh and OC43 Spike IgG. R values are determined using Spearman’s correlation and statistical significant correlations displayed as ***p<0.001. Dotted lines indicate the limit of detection following subtraction of DMSO from T cell response.

The coordinated recruitment of circulating Tfh for germinal centre reactions and early antibody production by plasmablasts is associated with seroconversion (Pilkinton et al., 2017). The early activated (ICOS^+^ PD-1^+^) total Tfh response was significantly increased in infected children compared to adults and negative controls (Supplementary Figure 4G), whilst plasmablast responses were increased in both children and adults compared to negative controls showing B cell recruitment with infection (Supplementary Figure 4H).

### T cell responses increase with time post infection and age

Longitudinal sample collection up to 180 days post infection enabled us to determine the trend of T cell responses with time post infection. Long-term stability of durable T cell immunity is likely important to minimise the symptom severity of reinfection with SARS-CoV-2. The CD4^+^ T cell response to structural peptides had stable responses post infection (Figure 2A) (r=0.1475, p=0.2265), whilst structure specific CD8^+^ T cell responses had a moderate significant trend for increased responses with time (Figure 2B) (r=0.4194, p=0.0003). This was also reflected in the acute fold changes of CD4^+^ and CD8^+^ T cell responses (Figure 1F), which indicates the CD4^+^ T cell response is recruited early during SARS-CoV-2 infection (Figure 1D), the CD8^+^ T cell response takes more time to build up with time post-infection. Furthermore, there was no difference in T cell exhaustion (by PD-1 expression) between infected adults and children at either acute or memory responses (Supplementary Figure 5).

The fold change of response magnitude for paired acute responses (<14 days) to memory time-points (>14 days) (Figure 2CD), showed comparable fold-changes in children and adults for CD4^+^ or CD8^+^ T cell response to most viral proteins. Only accessory-specific CD8^+^ T cell responses had a significant decrease in infected children (Figure 2D). Whilst the acute structural specific CD4^+^ T cell response was significantly increased in adults compared to negative controls (Figure 2E), the memory CD4^+^ and CD8^+^ T cell response were significantly lower in children compared to infected adults (Figure 2EH), resulting in a trend for significantly increased T cell responses with age (Figure 2FIJ), excluding acute CD8^+^ T cell responses (Figure 2G).

The difference in magnitude of T cell responses with age and time indicates functional differences in T cell recruitment and differentiation, therefore we assessed cytokine polyfunctionality and memory phenotypes. Cytokine polyfunctionality is associated with increased protection from infection for multiple viruses (Lichterfeld et al., 2004, Sridhar et al., 2013), and associated with cellular division and terminal differentiation (Denton et al., 2011). Whilst differentiation of T cell memory phenotypes occurs early during infection and can reflect the extent of inflammation (Kretschmer et al., 2020), impacting recall capacity long-term (Kinjyo et al., 2015) to infected tissues (Weninger et al., 2001).

Cytokine polyfunctionality of structure-specific T cells (Figure 3AB) was comparable between adults and children at acute (<14 days), convalescent (15-60 days) or memory (61-180 days) time points (Figure 3C), therefore on a per cell basis adults and children had comparable cytokine responses. The phenotype of structure-specific T cells at memory time points (Figure 3D), however showed that children had reduced T effector memory (TEM) CD4^+^ T cells compared to infected adults (Figure 3E). The phenotype of structure-specific CD8^+^ T cells was comparable (Figure 3F).

### Prior common cold coronavirus immunity and cellular responses

The level of coronavirus Spike-specific IgG was determined at early time points (<14 days) of SARS-CoV-2 infection, to determine if pre-existing immunity impacted T cell responses. The magnitude of α-coronavirus 229E and NL63-specific IgG was comparable between infected children and adults and adult negative controls (Figure 4A), whilst β-coronavirus HKU-1 and OC43-specific IgG was significantly lower in infected children than infected adults (Figure 4B). Furthermore, there was no difference in OC43-IgG responses between symptomatic or asymptomatic infections, with significance only being seen between symptomatic adults and children (Figure 4C). There was a significant correlation of OC43-

IgG responses with age (Figure 4D) (r=0.6466, p=0.0002). However there was no direct significant correlation between OC43-IgG responses and structure-specific CD4^+^ (p=0.1027, Figure 4E) or CD8^+^ T cells (p=0.9729, Figure 4F). Similar trends of a lack of significant correlation were seen for HKU-1 IgG responses and acute CD4^+^ T cell responses (r=0.3085, p=0.1034, *data not shown*), despite other reports in uninfected adults (Tan et al., 2021). However, there was a borderline moderate negative correlation between OC43-IgG and early acute activated Tfh responses (Figure 4G) (r=-0.3310, p=0.0854).

## Discussion

SARS-CoV-2 infection of children is associated with milder clinical outcomes than adults, and the immune mechanisms are unknown. Several immune mechanism have been proposed to explain these differences such as innate cell recruitment and impairment by autoantibodies (Wang et al., 2020), mobilisation of antibody responses, differing levels of pre-existing cross-reactive immunity by common cold coronavirus exposure (Anderson et al., 2020) or baseline total IgM levels (Selva et al., 2020). However, the SARS-CoV-2 T cell compartment in children has so far been under studied (Tosif et al., 2020). Viral loads (Jones et al., 2020) and neutralising antibody titers (Weisberg et al., 2021, Lau et al., 2021) are reportedly comparable when age is accounted for, however data is more limited in children. Viral loads, neutralising antibody titers (Garcia-Beltran et al., 2020), and T cell responses (Peng et al., 2020) impact clinical severity of COVID-19.

Cross-reactive T cell responses in unexposed adults have been mapped to have been NSPs of ORF1ab and Spike (Grifoni et al., 2020), whilst recent infection boosts structural Spike and N specific T cells (Le Bert et al., 2020, Mateus et al., 2020). The specificity of SARS-CoV-2 antibody landscapes differs in infected children to adults (Weisberg et al., 2021, Hachim et al., 2021), with an increased contribution by accessory proteins (Hachim et al., 2021), whilst the ORF1ab response is under characterised. SARS-CoV-2 antibody landscapes indicate that the specificity and balance of the adaptive immune responses in children is different to adults. We sought to determine the balance of SARS-CoV-2 specific T cell responses in adults and children as infection progresses to recovery for long-term memory, whilst considering T cell specificity for the virion (structural proteins) and virus replication (ORF1ab and accessory proteins) as a surrogate for viral replication and pathogenesis.

Overall, we found total IFNγ CD4^+^ and CD8^+^ T cell responses are significantly lower in SARS-CoV-2 infected children than adults against the viral structural proteins, and in CD8^+^ T cells against ORF1ab proteins. Whilst there were no negative control PBMCs available in our study from uninfected children for direct comparison as it is difficult to obtain blood from healthy children, pairwise comparison between infected adults and children showed markedly different SARS-CoV-2 T cell responses. However, we and others (Lewis et al., 1991, Chipeta et al., 1998, Rudolph et al., 2018, Booth et al., 2014), found that the capacity of children’s T cells to respond to polyclonal non-specific activation is also lower. Whereas vaccination with live influenza vaccines boosts T cell responses in children and not adults (He et al., 2006), this may be due to differences in prior immunity through infection (Shannon et al., 2019), resulting in qualitative differences in antigen experienced CD4^+^ T cell responses in children. As IFNγ T cell induction increases with age, so does exposure to viruses. Therefore the differences in SARS-CoV-2 T cell magnitude may be due to inherent differences between children and adult T cell threshold for activation. This has different consequences in different scenarios of a pandemic virus compared to vaccination building on prior memory. For example, there is an increased fold change of children’s T cell responses compared to adults during live attenuated influenza vaccination (He et al., 2006), whilst we found lower SARS-CoV-2 T cell response magnitudes in children, their fold changes and polyfunctional cytokines of the T cell responses was comparable between adults and children. Therefore there is equal recruitment of SARS-CoV-2 T cell responses in adults and children but likely different baseline levels of cross-reactive responses to recruit from and inherent differences in thresholds of activation.

A difference in T cell memory phenotypes showed greater bias towards T cell effector memory in adults compared to children. Along with total response and polyfunctional cytokine production, this indicates that on a per T cell basis the T cell response in children is less antigen experienced and matured than adults, and may be due to different levels of prior immunity to seasonal human coronaviruses. This was also found by smaller magnitude memory T cell responses in children than adults, which may imply a weaker long-term memory response in children potentially impacting outcomes at reinfection. Indeed, we found significantly lower levels of β-coronavirus specific antibodies in infected children than adults, and there was a significant trend for both increased SARS-CoV-2 specific T cell responses and OC43-specific IgG with increasing age. Recently, similar results were found in healthy adults as HKU-1 IgG showed an increasing trend with SARS-CoV-2 specific T cell responses of memory phenotype in uninfected adults (Tan et al., 2021). A borderline trend for decreasing acute activated Tfh with higher OC43-specific IgG levels also suggests a greater importance CD4^+^ T cell recruitment in more immunologically naïve settings, and as β-coronavirus specific IgG levels increase there is a decreasing drive for Tfh recruitment. Only the quantification of baseline T cell responses specific for common cold viruses and subsequent exposure to SARS-CoV-2 in further studies, such as in human cohort transmission settings or animal models, will determine if prior β-coronavirus immunity, based on T or B cells, has a protective role in COVID-19.

The quality of T cell responses, assessed by SARS-CoV-2 specific T cell polyfunctional cytokine production, was equivalent between children and adults, reflecting division and terminal differentiation. Furthermore, T cell exhaustion, assessed by expression of PD-1 is higher in COVID-19 patients with expression increasing with severity (Diao et al., 2020). While we also see differences in infected and negative adults, infected adults and children have equivalent PD-1 levels on T cells. Therefore, whilst SARS-CoV-2 specific T cell responses in children are reduced they comparably activated/exhausted (by PD-1 expression). The matched quality of response but higher threshold for IFNγ production by T cells in children may drive a less inflammatory environment that promotes more mild outcomes in children. Lower levels of total and inflammatory monocytes, may further contribute to a less inflammatory environment. However paradoxically lower inflammatory monocytes are associated with both healthy individuals and patients with severe COVID-19 infections (Mann el al., 2020; Zhang et al., 2021), therefore the timing of monocyte recruitment is likely important. There was a different recruitment of innate and adaptive cellular responses in adults and children during SARS-CoV-2 infection. Children had increased Tfh recruitment, comparable plasmablast responses, but reduced monocytes, specific CD4^+^ and CD8^+^ T cell responses, in both magnitude and proportion of responders. The inflammatory milieu is likely drivers of innate and adaptive cell recruitment, and indicates differences between adults and children across the anti-viral immune response.

T cell responses can be quantified by numerous methods besides IFNγ induction by peptide stimulation as adopted in our study, such as proliferation (Ki67) and activation induced markers (AIM, such as 41-BB/CD40L, OX40, CD25 etc) (Grifoni et al., 2020), antigen experience (CD69, CD44), or the production Th2 cytokines such as IL-4. In addition to functional assays, T cell responses can be quantified by T cell receptor binding to cognate peptide MHC by dimer/tetramers (Peng et al., 2020). However, known SARS-CoV-2 epitopes are currently limited and the HLA should be defined within donors. The induction of AIM for Tfh responses and the CD4^+^ T cell profile across different cell types (Th17, Treg etc), should also be assessed in future studies. We found that children had increased activated Tfh responses but lower IFNγ^+^ CD4^+^ T cells, therefore the CD4^+^ T cell compartment is modulated by SARS-CoV-2 infection, and more so than the CD8^+^ T cell response which is lagging behind the acute stage. In addition more variables can be assessed by high dimensional flow cytometry or RNAseq approaches, and further mechanistic studies are needed to define the basis of immunological differences between T cell responses of children and adults indicated in our study.

Adult negative controls had significantly lower structural/ORF1ab CD4^+^ T cell responses but comparable CD8^+^ T cell responses to infected adults. This was also reflected in greater CD4^+^ T cell increases at acute timepoints of infection, and delayed CD8^+^ T cell increases with time, indicating CD4^+^ T cell responses play a greater role in the early responses to SARS-CoV-2 infection. The contribution of different virion structural and non-structural proteins reflects MHC processing access during viral replication, whereby MHCII access to structural proteins elicited substantial CD4^+^ T cell responses in adults, in children the CD4^+^ T cell response was predominantly ORF1ab specific. The imbalance of peptide specificities for non-structural proteins for children’s CD4^+^ T cell compartment may indicate either different virus replication and pathogenesis at the cellular level or incomplete recruitment of *de novo* CD4^+^ T cell responses. Previously, in MERS-CoV infection, the magnitude of the CD4^+^ T cell response is proportional to virus replication and duration of illness (Zhao et al., 2017). This is consistent with the mild outcomes of COVID-19 in children and reduced T cell responses reported here in our study of mild and asymptomatic SARS-CoV-2 infection. We cannot attribute the differences in T cell response magnitude with severity of illness in children to adults, unlike others reports (Peng et al., 2020), as the majority of both infections we studied are mild or asymptomatic. Therefore children have reduced SARS-CoV-2 T cell responses due to lower baseline immune activation, and further research is still needed to discern the protective role of T cells in COVID-19.

## Acknowledgements

This study was partly supported by the Theme based Research Grants Scheme (T11-712/19-N), Health and Medical Research Fund (HMRF COVID- 190115 and COVID-190126), National Institutes of Allergy and Infectious Diseases, National Institutes of Health (USA) (HHSN272201400006C and U01AI151810). We thank Daniela Weiskopf, Jose Barrera, Shane Crotty and Alessandro Sette from La Jolla Institute, USA, for providing SARS-CoV-2 peptide Megapools. This project utilised an Invitrogen Attune flow cytometer assisted by the Pasteur Foundation Asia.

## Author contributions

CAC performed experiments. CAC, APYL, AH, NK, SAV designed experiments. MYWK, WHC, YSY, SSC OTYT, DSCH provided clinical information and supplied clinical samples, processed by FNLM coordinated by SMSC. EHYL performed further analysis. CAC, LP, JSMP and SAV designed the study. CAC, APYL, AH, NK, LP, JSMP, SAV interpreted results and wrote the manuscript.

