## Supplementary figures and tables for "SARS-CoV-2 specific T cell responses are lower in children and increase with age and time after infection"

**Supplementary Table 1.** T cell response in COVID-19 patients, trend and versus controls

| | | Trend days after symptom onset | | Children vs Adults | | COVID-19 cases vs Controls | | |
|---|---|---|---|---|---|---|---|---|
| **T cells** | | Estimate | P-value | Estimate | P-value | Median (case) | Median (negative) | P-value |
| **CD4** | Structural IFNγ$^{+}$ (total) | 0.000012 | 0.920 | -0.029 | 0.01 | 0.0479 | 0.00999 | 0.003 |
| | ORF1ab IFNγ$^{+}$ (total) | 0.00016 | 0.310 | 0.017 | 0.297 | 0.0235 | 0.0115 | 0.004 |
| | Accessory IFNγ$^{+}$ (total) | -0.000037 | 0.256 | -0.0043 | 0.153 | 0.0124 | 0.0134 | 0.350 |
| | Structural TEM | 0.0032 | 0.950 | -3.2 | 0.493 | 88.7 | 89.5 | 0.552 |
| | Structural TCM | -0.017 | 0.577 | 1.5 | 0.625 | 9.28 | 8.54 | 0.752 |
| | Structural IFNγ$^{+}$TNFα$^{+}$ | -0.0000095 | 0.637 | -6.00E-04 | 0.756 | 0.00436 | 0.000813 | 0.051 |
| | Structural IFNγ$^{+}$TNFα$^{+}$IL2$^{+}$ | -0.000086 | 0.003 | -0.0077 | 0.232 | 0.00342 | 0.000723 | 0.068 |
| **CD8** | Structural IFNγ$^{+}$ (total) | 0.00014 | 0.592 | -0.094 | 0.009 | 0.0345 | 0.0395 | 0.884 |
| | ORF1ab IFNγ$^{+}$ (total) | 0.00052 | 0.596 | -0.27 | 0.008 | 0.101 | 0.101 | 0.818 |
| | Accessory IFNγ$^{+}$ (total) | -0.000029 | 0.084 | -0.0021 | 0.184 | 0.00796 | 0.0157 | 0.156 |
| | Structural TEM | -0.045 | 0.452 | 7.7 | 0.232 | 77.4 | 68.5 | 0.350 |
| | Structural TCM | 0.10 | 0.006 | -2.1 | 0.546 | 10.6 | 20.6 | 0.022 |
| | Structural IFNγ$^{+}$TNFα$^{+}$ | 0.0000050 | 0.678 | -0.0015 | 0.219 | 0.000595 | 0 | 0.036 |
| | Structural IFNγ$^{+}$TNFα$^{+}$IL2$^{+}$ | 0.00000031 | 0.845 | -0.00041 | 0.276 | 0 | 0 | 0.315 |

* P value by Mann-Whitney test, significant values in red

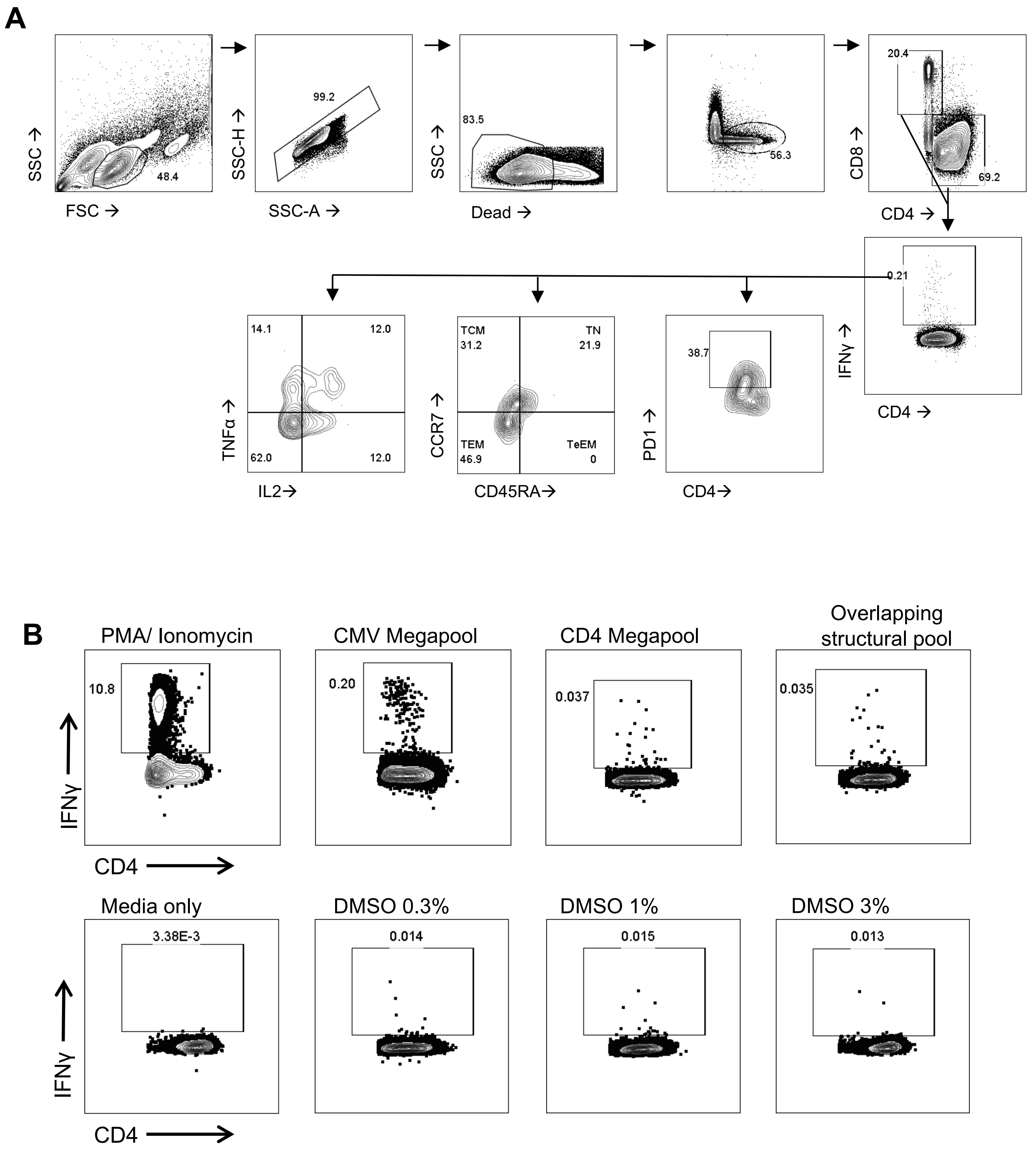


**Supplementary Figure 1 – SARS-CoV-2 specific cells for cytokine production and phenotype by Flow cytometry.** (A) The gating strategy for the characterisation of IFNγ$^{+}$ responses for CD4$^{+}$ and CD8$^{+}$ T cells for cytokine production (IFNγ, TNFa, IL-2), memory phenotype (CCR7, CD45RA), and exhaustion markers (PD-1). (B) FACS plots showing CD4$^{+}$ IFNγ production to positive controls, PMA/Ionomycin, a CMV megapool and a CD4 specific megapool (from Grifoni *et al*. 2020), and negative controls with a range of DMSO concentrations.

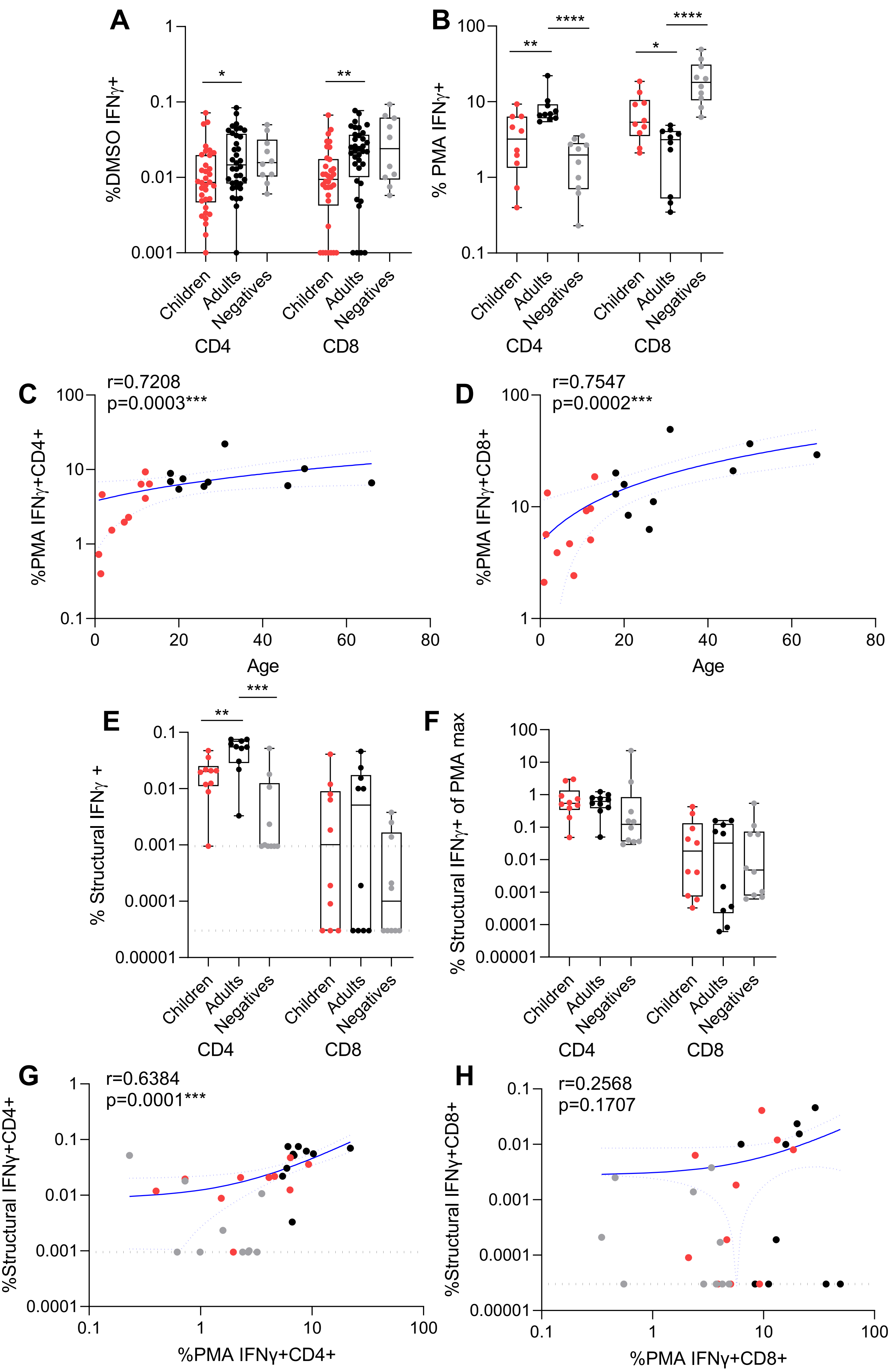


**Supplementary Figure 2**

**Supplementary Figure 2 – Maximum $CD4^+$ and $CD8^+$ IFNγ T cell responses increase with age.** $CD4^+$ and $CD8^+$ T cell responses for (A) background (by DMSO stimulation) and (B) maximum (by PMA/Ionomycin stimulation) in children (n=10) and adults (n=10) from convalescent/memory time points (mean±stdev: 56.9±29.7, range: 27-138 days post symptom onset), and uninfected adult controls (n=10). Comparisons by Mann Whitney test where $^{**}p<0.01$, $^{***}p<0.001$, $^{****}p<0.0001$. Correlation of age with $CD4^+$ (C) and $CD8^+$ T cell responses by PMA/ionomycin stimulation. Spearman's test was used to calculate r values, and statistical significance is displayed as $^{***}p<0.001$. Blue lines of linear regression represent the overall trend with dotted lines showing 95% confidence intervals. Black dotted lines represent the limit of detection (IFNγ of $CD4^+$=0.0095%, IFNγ of $CD8^+$=0.00003%). (E) The structural peptide pool response for $CD4^+$ and $CD8^+$ T cells in adults and children (F) normalised to a paired maximum IFNγ production from (B) PMA/ionomycin stimulation. Correlation of IFNγ production against structural pool and PMA/Ionomycin in $CD4^+$(G) and $CD8^+$ T cells cells (H).

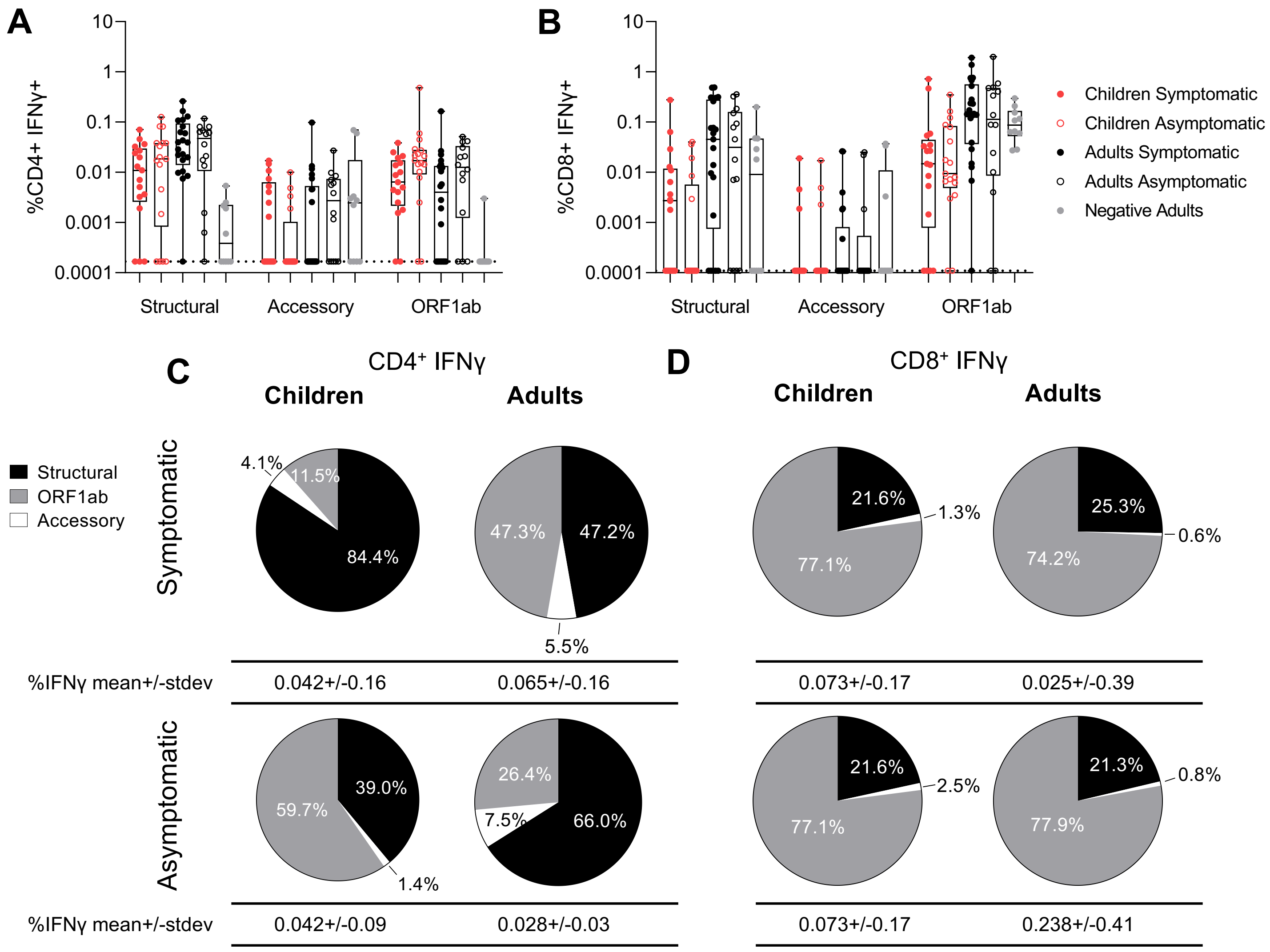


**Supplementary Figure 3 - IFNγ CD4⁺ and CD8⁺ T cell responses are not different between symptomatic or asymptomatic SARS-CoV-2 infected children or adults.** The SARS-CoV-2 CD4⁺ (A) or CD8⁺ (B) T cell responses of COVID-19 symptomatic children (n= 17, mean±stdev: 38.8±40.6 days), asymptomatic children (n= 17, 26.2±39.3 days), symptomatic adults (n= 22, 60.3±55.5 days), asymptomatic adults (n= 14, 32.8±32.3 days), from acute, convalescent and long-term memory time points (day 1 to 180 post symptom onset) and negative controls (n=10). Data represents the individual response with background subtracted, mean with upper and lower quartiles, minimum and maximum. Multiple comparisons were performed using Kruskal-Wallis test for between-group comparison. Pie charts of total IFNγ⁺ CD4⁺ (C) and CD8⁺ (D) T cell SARS-CoV-2 responses with background subtracted and non-responders assigned a response of zero (from A, B).

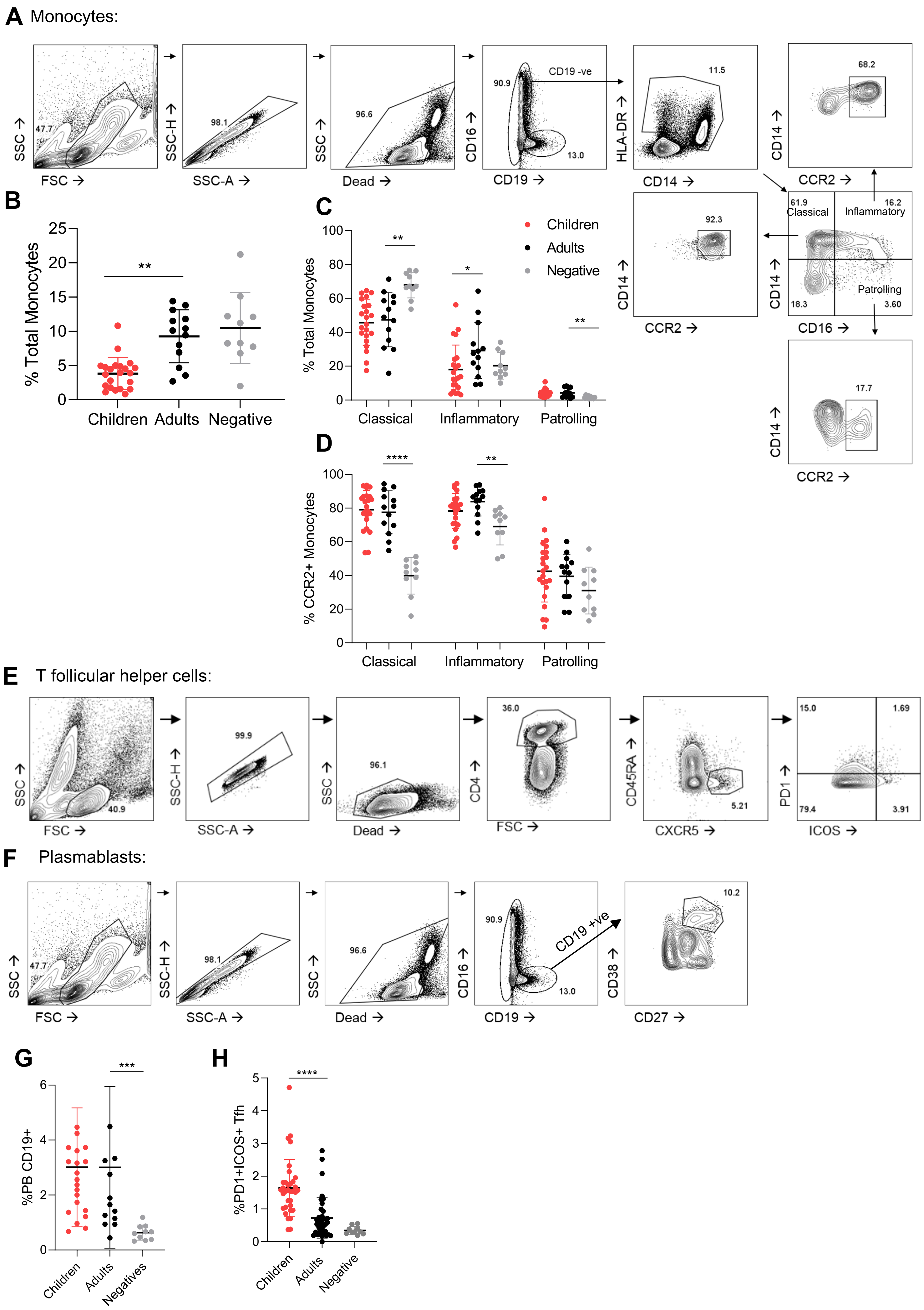


**Supplementary Figure 4**

**Supplementary Figure 4 – Cellular recruitment during acute infection of children shows increased Tfh responses.** (A) The gating strategy for characterisation of total, classical, inflammatory and patrolling monocytes and their activation levels (by CCR2). Early (<day 14) recruitment of innate and adaptive cells was measured by flow cytometry for COVID-19 children (n= 22), adults (n= 13) and negative controls (n=10). The total monocytes (B), monocyte phenotype (C), and activation of monocytes (D). The T follicular helper (E) and plasmablast (F) response was determined for activated Tfh (G), and total plasmablast (H). Data represents the individual response, mean±SD. Statistical differences were determined using Mann-Whitney test between children and adults, or adults and negatives where $^{**}p<0.01$, $^{***}p<0.001$, $^{****}p<0.0001$.

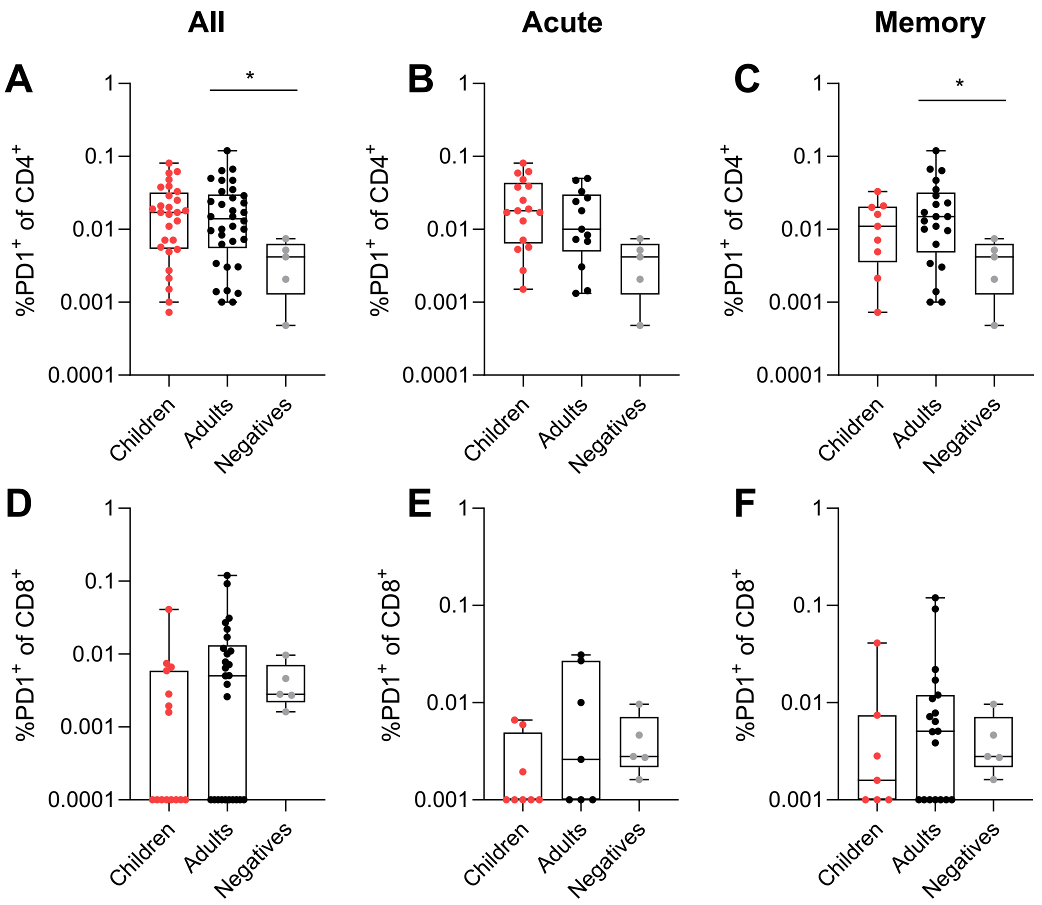


**Supplementary Figure 5 – T cell exhaustion is equivalent in adult and children CD4 and CD8 T cells.** PD1+ IFNγ+ CD4 (A-C) and CD8 (D-F) T cells in response to stimulation with structural pool at all time points (A, D) (day 1-180) in children (red), adults (black) and negative adults (grey) who are responders. Data separated into acute (B, E) (day<14 post symptom onset) and convalescent/memory (C, F) (day 15-180 post infection) time points. Comparisons between infected children and adults, or infected adults and negative adults are carried out using Mann Whitney (unpaired) test where $*p<0.05$.
